# Protection of natural infection against reinfection with SARS-CoV-2 JN.1 variant

**DOI:** 10.1101/2024.02.22.24303193

**Authors:** Hiam Chemaitelly, Peter Coyle, Mohamed Ali Ben Kacem, Houssein H. Ayoub, Patrick Tang, Mohammad R. Hasan, Hadi M. Yassine, Asmaa A. Al Thani, Zaina Al-Kanaani, Einas Al-Kuwari, Andrew Jeremijenko, Anvar H. Kaleeckal, Ali N. Latif, Riyazuddin M. Shaik, Hanan F. Abdul-Rahim, Gheyath K. Nasrallah, Mohamed Ghaith Al-Kuwari, Adeel A. Butt, Hamad E. Al-Romaihi, Mohamed H. Al-Thani, Abdullatif Al-Khal, Roberto Bertollini, Laith J. Abu-Raddad

## Abstract

This study investigated the effectiveness of natural infection in preventing reinfection with the JN.1 variant during a large JN.1 wave in Qatar, using a test-negative case-control study design. The overall effectiveness of previous infection in preventing reinfection with JN.1 was estimated at only 1.8% (95% CI: −9.3-12.6%). This effectiveness demonstrated a rapid decline over time since the previous infection, decreasing from 82.4% (95% CI: 40.9-94.7%) within 3 to less than 6 months after the previous infection to 50.9% (95% CI: −11.8-78.7%) in the subsequent 3 months, and further dropping to 18.3% (95% CI: −34.6-56.3%) in the subsequent 3 months. Ultimately, it reached a negligible level after one year. The findings show that the protection of natural infection against reinfection with JN.1 is strong only among those who were infected within the last 6 months, with variants such as XBB*. However, this protection wanes rapidly and is entirely lost one year after the previous infection. The findings support considerable immune evasion by JN.1.

## Main text

Evidence at the level of neutralizing antibodies suggests that the SARS-CoV-2 JN.1 variant demonstrates increased immune evasion compared to its parent lineage BA.2.86 and to recently circulating variants, such as XBB.1.5 and EG.5.1.^1^ JN.1 has also exhibited a growth advantage over other variants and triggered large SARS-CoV-2 waves in various countries,^2^ prompting the World Health Organization to classify it as a variant of interest on December 19, 2023.^2^ We estimated the effectiveness of natural infection in preventing reinfection with JN.1 during a large JN.1 wave in Qatar using the test-negative case-control study design.^3,4^

Qatar’s national COVID-19 databases were analyzed between December 4, 2023, when JN.1 dominated incidence (Figure S1 of the Supplementary Appendix), and February 12, 2024. These databases encompass all laboratory and medically supervised SARS-CoV-2 testing, infection clinical outcomes, COVID-19 vaccination, and demographic details within the country (Sections S1-S2).

Cases (SARS-CoV-2-positive tests) and controls (SARS-CoV-2-negative tests) were matched exactly one-to-two by factors that could influence the risk of infection, including sex, 10-year age group, nationality, number of coexisting conditions, number of vaccine doses, calendar week of the SARS-CoV-2 test, method of testing (polymerase chain reaction versus rapid antigen), and reason for testing (Section S3). Previous infection was defined as a SARS-CoV-2-positive test ≥90 days before the study test. Subgroup analyses estimating effectiveness against specifically symptomatic reinfection, and by vaccination status, were conducted.

Figure S2 and Table S1, respectively, show the study population selection process and characteristics. The study population was broadly representative of Qatar’s population. The overall effectiveness of previous infection in preventing reinfection with JN.1, regardless of symptoms, was estimated at 1.8% (95% CI: −9.3-12.6%) (Figure 1). This effectiveness demonstrated a rapid decline over time since the previous infection, decreasing from 82.4% (95% CI: 40.9-94.7%) within 3 to less than 6 months after the previous infection to 50.9% (95% CI: −11.8-78.7%) in the subsequent 3 months, and further dropping to 18.3% (95% CI: −34.6-56.3) in the subsequent 3 months. Ultimately, it reached a negligible level after one year. The effectiveness was estimated at 49.1% (95% CI: 20.4-67.5%) during the first year and at −2.5% (95% CI: −13.5-9.0%) thereafter.

**Figure 1.**
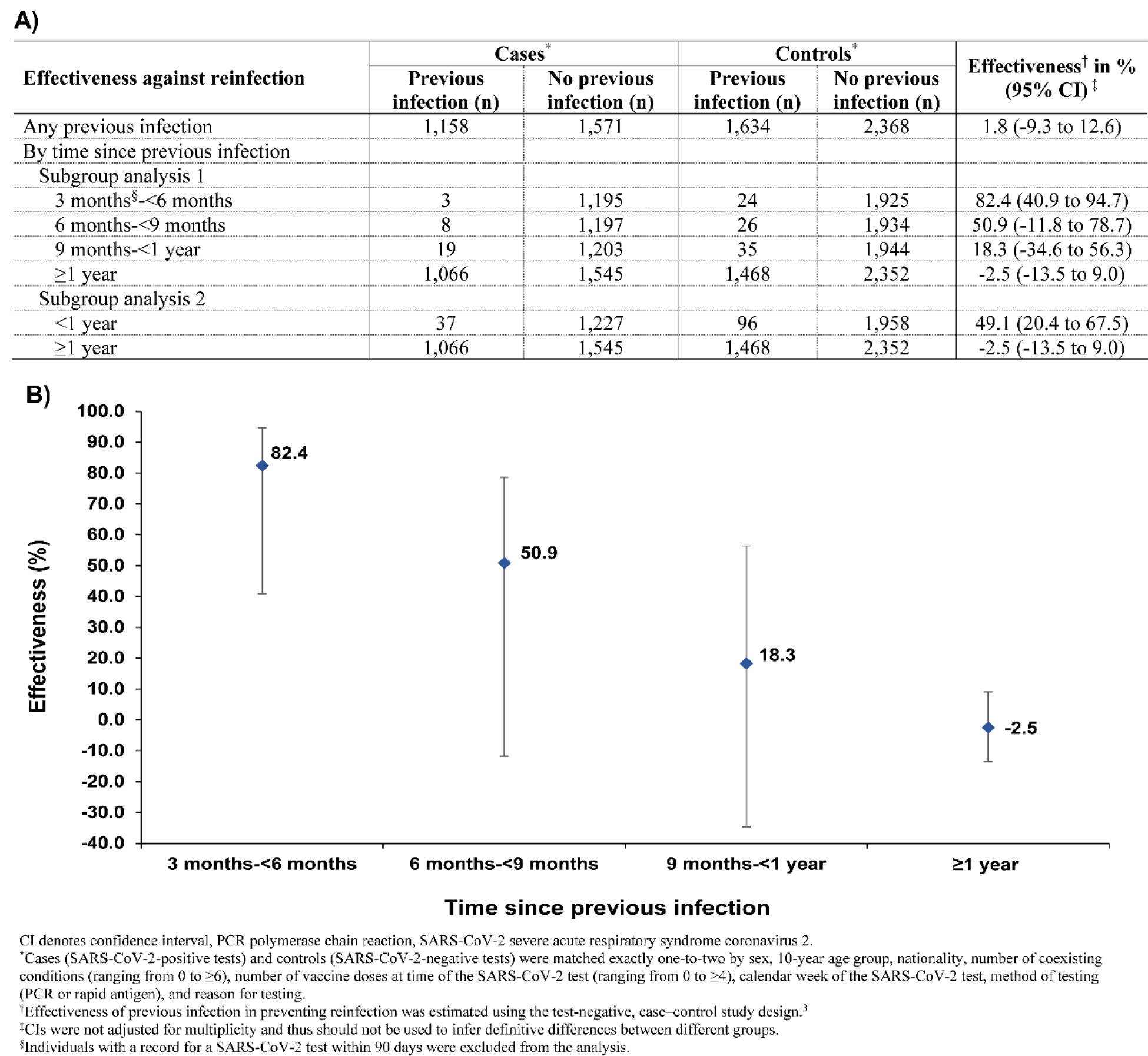
Protection against reinfection with JN.1, irrespective of symptoms, overall (A) and by time since previous infection (A and B)

The effectiveness against symptomatic reinfection with JN.1 demonstrated a similar pattern to that observed for any reinfection (Table S2). The overall effectiveness against symptomatic reinfection was −2.3% (95% CI: −14.4-10.3%). Subgroup analyses for unvaccinated and vaccinated individuals yielded results similar to those of the main analysis (Table S2). Limitations are discussed in Section S3.

The protection of natural infection against reinfection was strong among those who were infected within the last 6 months, with variants such as XBB*. However, this protection waned rapidly and was entirely lost one year after the previous infection. These findings support a considerable immune evasion by JN.1, and that this immune evasion led to the observed rapid waning of the protection against JN.1 (Figure 1), a pattern for the effect of immune evasion first characterized for SARS-CoV-2 following the omicron variant emergence at the end of 2021.^4,5^

### Oversight

The institutional review boards at Hamad Medical Corporation and Weill Cornell Medicine– Qatar approved this retrospective study with a waiver of informed consent. The study was reported according to the Strengthening the Reporting of Observational Studies in Epidemiology (STROBE) guidelines (Table S3). The authors vouch for the accuracy and completeness of the data and for the fidelity of the study to the protocol. Data used in this study are the property of the Ministry of Public Health of Qatar and were provided to the researchers through a restricted-access agreement for preservation of confidentiality of patient data. The funders had no role in the study design; the collection, analysis, or interpretation of the data; or the writing of the manuscript.

### Author contributions

HC co-designed the study, performed the statistical analyses, and co-wrote the first draft of the article. LJA conceived and co-designed the study, led the statistical analyses, and co-wrote the first draft of the article. HC and LJA accessed and verified all the data. PVC conducted viral genome sequencing and designed mass PCR testing to allow routine capture of variants. MBK conducted viral genome sequencing. PT and MRH designed and conducted multiplex, RT-qPCR variant screening and viral genome sequencing. HMY and AAAT conducted viral genome sequencing. All authors contributed to data collection and acquisition, database development, discussion and interpretation of the results, and to the writing of the article. All authors have read and approved the final manuscript.

## Data Availability

The dataset of this study is a property of the Qatar Ministry of Public Health that was provided to the researchers through a restricted access agreement that prevents sharing the dataset with a third party or publicly. Future access to this dataset can be considered through a direct application for data access to Her Excellency the Minister of Public Health
(https://www.moph.gov.qa/english/Pages/default.aspx). Aggregate data are available within the manuscript and its Supplementary information.

## Acknowledgements and support

We acknowledge the many dedicated individuals at Hamad Medical Corporation, the Ministry of Public Health, the Primary Health Care Corporation, Qatar Biobank, Sidra Medicine, and Weill Cornell Medicine-Qatar for their diligent efforts and contributions to make this study possible.

The authors are grateful for institutional salary support from the Biomedical Research Program and the Biostatistics, Epidemiology, and Biomathematics Research Core, both at Weill Cornell Medicine-Qatar, as well as for institutional salary support provided by the Ministry of Public Health, Hamad Medical Corporation, and Sidra Medicine. The authors are also grateful for the Qatar Genome Programme and Qatar University Biomedical Research Center for institutional support for the reagents needed for the viral genome sequencing. The funders of the study had no role in study design, data collection, data analysis, data interpretation, or writing of the article. Statements made herein are solely the responsibility of the authors.

## Supplementary Appendix

### Acknowledgments

The authors are grateful for institutional salary support from the Biomedical Research Program and the Biostatistics, Epidemiology, and Biomathematics Research Core, both at Weill Cornell Medicine-Qatar, as well as for institutional salary support provided by the Ministry of Public Health, Hamad Medical Corporation, and Sidra Medicine. The authors are also grateful for the Qatar Genome Programme and Qatar University Biomedical Research Center for institutional support for the reagents needed for the viral genome sequencing. The funders of the study had no role in study design, data collection, data analysis, data interpretation, or writing of the article.

Statements made herein are solely the responsibility of the authors.

### Author contributions

**Figure S1.**
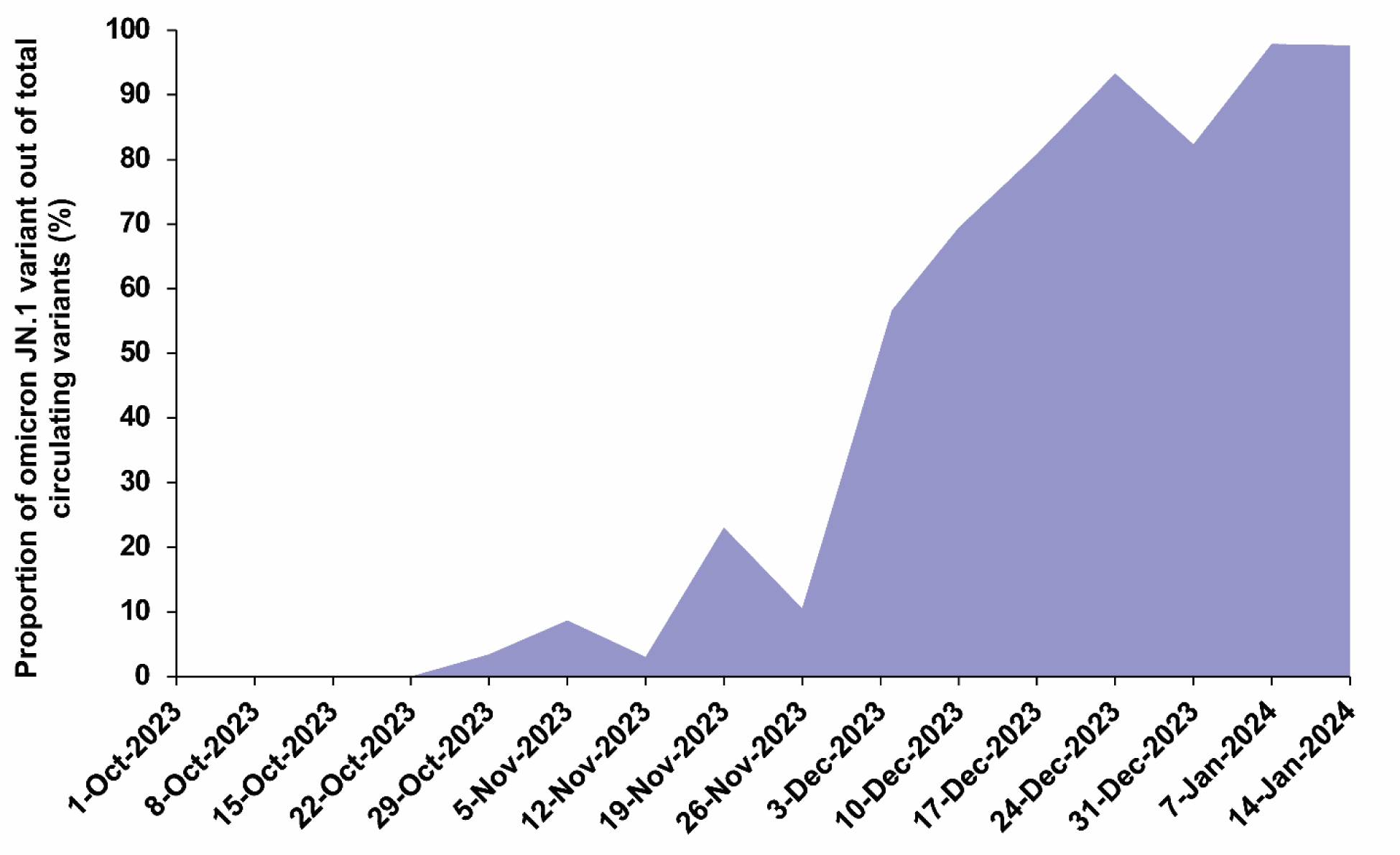
Qatar’s SARS-CoV-2 variant genomic surveillance. Results of viral genome sequencing of 515 samples between October 1, 2023 and January 20, 2024, illustrating the dominance of the omicron JN.1 variant starting from early December 2023.

### Section S1. Data sources

Qatar’s national and universal public healthcare system uses the Cerner-system advanced digital health platform to track all electronic health record encounters of each individual in the country, including all citizens and residents registered in the national and universal public healthcare system. Registration in the public healthcare system is mandatory for citizens and residents.

The databases analyzed in this study are data-extract downloads from the Cerner-system that have been implemented on a regular schedule since the onset of the pandemic by the Business Intelligence Unit at Hamad Medical Corporation. Hamad Medical Corporation is the national public healthcare provider in Qatar. At every download all tests, coronavirus disease 2019 (COVID-19) vaccinations, hospitalizations related to COVID-19, and all death records regardless of cause are provided to the authors through .csv files. These databases have been analyzed throughout the pandemic not only for study-related purposes, but also to provide policymakers with summary data and analytics to inform the national response.

Every health encounter in the Cerner-system is linked to a unique individual through the HMC Number that links all records for this individual at the national level. Databases were merged and analyzed using the HMC Number to link all records whether for testing, vaccinations, hospitalizations, and deaths. All COVID-19-related healthcare was provided only in the public healthcare system. No private entity was permitted to provide COVID-19-related hospitalization. COVID-19 vaccination was also provided only through the public healthcare system. These health records were tracked throughout the COVID-19 pandemic using the Cerner system. This system has been implemented in 2013, before the onset of the pandemic. Therefore, we had the health records related to this study for the full national cohort of citizens and residents throughout the pandemic.

Demographic details for every HMC Number (individual) such as sex, age, and nationality are collected upon issuing of the universal health card, based on the Qatar Identity Card, which is a mandatory requirement by the Ministry of Interior to every citizen and resident in the country. Data extraction from the Qatar Identity Card to the digital health platform is performed electronically through scanning techniques.

All severe acute respiratory syndrome coronavirus 2 (SARS-CoV-2) testing in any facility in Qatar is tracked nationally in one database, the national testing database. This database covers all testing in all locations and facilities throughout the country, whether public or private. Every polymerase chain reaction (PCR) test and a proportion of the facility-based rapid antigen tests conducted in Qatar, regardless of location or setting, are classified on the basis of symptoms and the reason for testing (clinical symptoms, contact tracing, surveys or random testing campaigns, individual requests, routine healthcare testing, pre-travel, at port of entry, or other).

Before November 1, 2022, SARS-CoV-2 testing in Qatar was done at a mass scale where about 5% of the population were tested every week.^1,2^ Based on the distribution of the reason for testing up to October 31, 2022, most of the tests in Qatar were conducted for routine reasons, such as being travel-related, and about 75% of cases were diagnosed not because of appearance of symptoms, but because of routine testing.^1,2^ Subsequently, testing rates decreased, with less than 1% of the population being tested per week.^1,3^All testing results in the national testing database during the present study were factored in the analyses of this study.

The first large omicron wave that peaked in January of 2022 was massive and strained the testing capacity in the country.^2–5^ Accordingly, rapid antigen testing was introduced to relieve the pressure on PCR testing. Implementation of this change in testing policy occurred quickly precluding incorporation of reason for testing in a large proportion of the rapid antigen tests. While the reason for testing is available for all PCR tests, it is not available for all rapid antigen tests. Availability of reason for testing for the rapid antigen tests also varied with time.

Rapid antigen test kits are available for purchase in pharmacies in Qatar, but outcome of home-based testing is not reported nor documented in the national databases. Since SARS-CoV-2-test outcomes were linked to specific public health measures, restrictions, and privileges, testing policy and guidelines stress facility-based testing as the core testing mechanism in the population. While facility-based testing is provided free of charge or at low subsidized costs, depending on the reason for testing, home-based rapid antigen testing is de-emphasized and not supported as part of national policy.

Qatar launched its COVID-19 vaccination program in December 2020, employing mRNA vaccines and prioritizing individuals based on coexisting conditions and age criteria.^1,6^ COVID-19 vaccination was provided free of charge, regardless of citizenship or residency status, and was nationally tracked.^1,6^

Coexisting conditions are ascertained and classified based on the ICD-10 codes for the conditions as recorded in the electronic health record encounters of each individual in the Cerner-system national database that includes all citizens and residents registered in the national and universal public healthcare system. The public healthcare system provides healthcare to the entire resident population of Qatar free of charge or at heavily subsidized costs, including prescription drugs. With the mass expansion of this sector in recent years, facilities have been built to cater to specific needs of subpopulations. For example, tens of facilities have been built, including clinics and hospitals, in localities with high density of craft and manual workers.^7^

All encounters for each individual are analyzed to determine the coexisting-condition classification for that individual, as part of a recent national analysis to assess healthcare needs and resource allocation. The Cerner-system national database includes encounters starting from 2013, after this system was launched in Qatar. As long as each individual had at least one encounter with a specific coexisting-condition diagnosis since 2013, this person was classified with this coexisting condition.

Individuals who have coexisting conditions but never sought care in the public healthcare system, or seek care exclusively in private healthcare facilities, were classified as individuals with no coexisting condition due to absence of recorded encounters for them.

Qatar has unusually young, diverse demographics, in that only 9% of its residents are ≥50 years of age, and 89% are expatriates from over 150 countries.^8,9^ Further descriptions of the study population and these national databases were reported previously.^1,2,5,9–13^

### Section S2. Laboratory methods and variant ascertainment

#### Real-time reverse-transcription polymerase chain reaction testing

Nasopharyngeal and/or oropharyngeal swabs were collected for PCR testing and placed in Universal Transport Medium (UTM). Aliquots of UTM were: 1) extracted on KingFisher Flex (Thermo Fisher Scientific, USA), MGISP-960 (MGI, China), or ExiPrep 96 Lite (Bioneer, South Korea) followed by testing with real-time reverse-transcription PCR (RT-qPCR) using TaqPath COVID-19 Combo Kits (Thermo Fisher Scientific, USA) on an ABI 7500 FAST (Thermo Fisher Scientific, USA); 2) tested directly on the Cepheid GeneXpert system using the Xpert Xpress SARS-CoV-2 (Cepheid, USA); or 3) loaded directly into a Roche cobas 6800 system and assayed with the cobas SARS-CoV-2 Test (Roche, Switzerland). The first assay targets the viral S, N, and ORF1ab gene regions. The second targets the viral N and E-gene regions, and the third targets the ORF1ab and E-gene regions.

All PCR testing was conducted at the Hamad Medical Corporation Central Laboratory or Sidra Medicine Laboratory, following standardized protocols.

#### Rapid antigen testing

SARS-CoV-2 antigen tests were performed on nasopharyngeal swabs using one of the following lateral flow antigen tests: Panbio COVID-19 Ag Rapid Test Device (Abbott, USA); SARS-CoV-2 Rapid Antigen Test (Roche, Switzerland); Standard Q COVID-19 Antigen Test (SD Biosensor, Korea); or CareStart COVID-19 Antigen Test (Access Bio, USA). All antigen tests were performed point-of-care according to each manufacturer’s instructions at public or private hospitals and clinics throughout Qatar with prior authorization and training by the Ministry of Public Health (MOPH). Antigen test results were electronically reported to the MOPH in real time using the Antigen Test Management System which is integrated with the national COVID-19 database.

#### Classification of infections by variant type

Surveillance for SARS-CoV-2 variants in Qatar is based on viral genome sequencing and multiplex RT-qPCR variant screening^14^ of weekly collected random positive clinical samples,^1,15–19^ complemented by deep sequencing of wastewater samples.^17,20,21^ Further details on the viral genome sequencing and multiplex RT-qPCR variant screening throughout the SARS-CoV-2 waves in Qatar can be found in previous publications.^1,2,4,11,15–19,22–26^

### Section S3. Detailed study methods

#### Study population and study design

The study was conducted on the population of Qatar between December 4, 2023, when JN.1 dominated incidence in Qatar (Figure S1), and February 12, 2024, marking the end of the study period. The effectiveness of natural infection against reinfection was estimated, both overall and by time since previous infection utilizing 3-month intervals, using the test-negative, case-control study design.^2,4,25–29^ This design compares the odds of previous infection among SARS-CoV-2-positive tests (cases) and SARS-CoV-2-negative tests (controls).^2,4,25–31^

Cases and controls were defined as SARS-CoV-2-positive and SARS-CoV-2-negative tests conducted during the analysis period, respectively. SARS-CoV-2 reinfection is conventionally defined as a documented infection ≥90 days after a previous infection, to avoid misclassifying prolonged test positivity as reinfection with shorter time intervals.^2,4,32,33^ Consequently, cases or controls preceded by SARS-CoV-2-positive tests within 90 days were excluded. To comply with the non-differential healthcare-seeking behavior assumption inherent to the test-negative study design,^27,30,31^ only tests with a documented reason for testing were included in the analysis.

Cases and controls were matched exactly one-to-two by sex, 10-year age group, nationality, number of coexisting conditions (ranging from 0 to ≥6), number of vaccine doses (ranging from 0 to ≥4), calendar week of the SARS-CoV-2 test, method of testing (PCR or rapid antigen), and reason for testing. This matching strategy aimed to balance observed confounders that could potentially influence the risk of infection across the exposure groups.^9,34–36^ The selection of matching factors was guided by findings from earlier studies on Qatar’s population.^1,6,16,26,37,38^

#### Statistical analysis

All records of SARS-CoV-2 testing were examined for the selection of cases and controls, but only matched samples were analyzed. Cases and controls were described using frequency distributions and measures of central tendency and compared using standardized mean differences (SMDs). An SMD of ≤0.1 indicated adequate matching.^39^

Odds ratios (ORs), comparing odds of previous infection among cases versus controls, and associated 95% confidence intervals (CIs) were derived using conditional logistic regression. Analyses stratified by time since previous infection considered the date for the most recent documented infection. CIs were not adjusted for multiplicity and interactions were not investigated. The reference group for all estimates comprised individuals with no documented previous infection.

Effectiveness measures and associated 95% CIs were calculated as 1-OR of previous infection among cases versus controls if the OR was <1, and as 1/OR-1 if the OR was ≥1.^5,27,30,40^ This approach ensured a symmetric scale for both negative and positive effectiveness, spanning from −100%-100%, resulting in a clear and meaningful interpretation of effectiveness, regardless of the value being positive or negative.

In addition to estimating the effectiveness of previous infection against any reinfection, whether asymptomatic or symptomatic, effectiveness was also assessed specifically against symptomatic reinfection. This was accomplished by restricting the analysis to tests performed due to clinical suspicion, indicating the presence of symptoms consistent with a respiratory tract infection. Subgroup analyses were performed, considering only unvaccinated and vaccinated individuals, respectively. Statistical analyses were conducted in STATA/SE version 18.0 (Stata Corporation, College Station, TX, USA).

#### Ethical approval

The institutional review boards at Hamad Medical Corporation and Weill Cornell Medicine– Qatar approved this retrospective study with a waiver of informed consent. The study was reported according to the Strengthening the Reporting of Observational Studies in Epidemiology (STROBE) guidelines (Table S4).

#### Limitations

The study is based on documented SARS-CoV-2 infections, but many infections may not have been documented, more so since the reduction in testing starting from November 1, 2022. However, this may not have appreciably affected our estimates, as it has been demonstrated that even substantial misclassification of previous infection status had a minimal impact on the estimated effectiveness of previous infection,^27^ a key strength of the test-negative design.^27^

A large proportion of SARS-CoV-2 tests lacked a specified reason for testing and were consequently excluded from the analysis. This led to a reduction in the sample size of the analyses and resulted in wide confidence intervals around some of the estimates.

With the relatively young population of Qatar,^9^ our findings may not be generalizable to other countries where elderly citizens constitute a large proportion of the population. While robust matching was implemented, the availability of data prevented matching on other factors such as geography or occupation. However, being essentially a city state, infection incidence in Qatar is distributed across neighborhoods. Nationality, age, and sex provide a powerful proxy for socio-economic status in this country,^9,34–36^ and thus matching by these factors may have also, at least partially, controlled for other factors such as occupation. This matching approach has been previously investigated in studies of different epidemiologic designs, and using control groups to test for null effects.^1,6,16,37,41^ These studies have supported that this matching prescription effectively controls for differences in infection exposure.^1,6,16,37,41^

However, bias in real-world data can arise unexpectedly or from unknown sources, such as subtle differences in test-seeking behavior, changes in testing patterns due to policy shifts, variations in test accessibility, or differences in the tendency to get tested between recoverees from previous infection and those who had not been infected or whose previous infection was undocumented.

**Figure S2.**
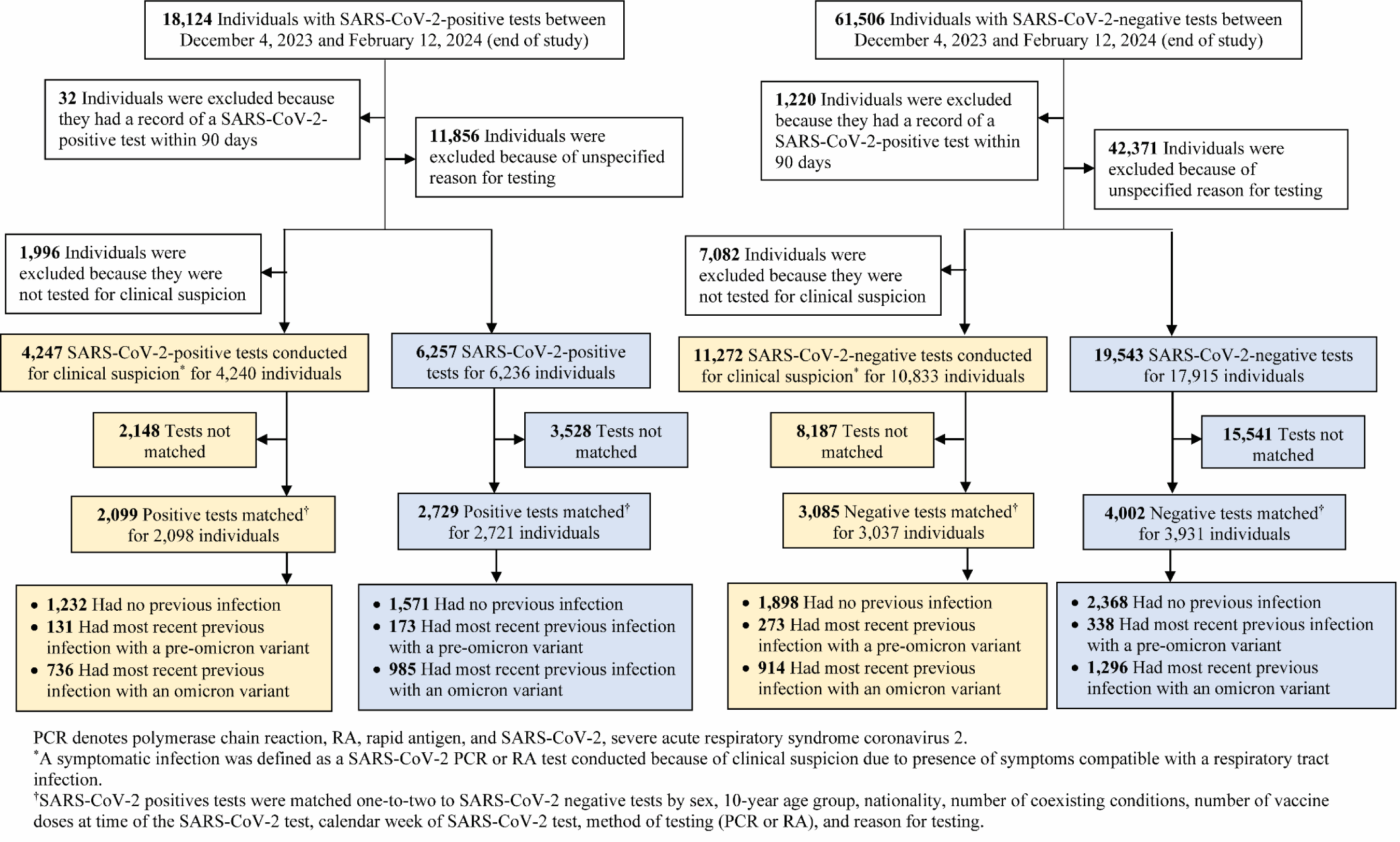
Flowchart describing the population selection process for investigating the effectiveness of natural infection in preventing reinfection with the omicron JN.1 variant. The blue color stands for the analysis for any reinfection regardless of symptoms. The yellow color stands for the analysis for specifically symptomatic reinfection.

**Table S1.**
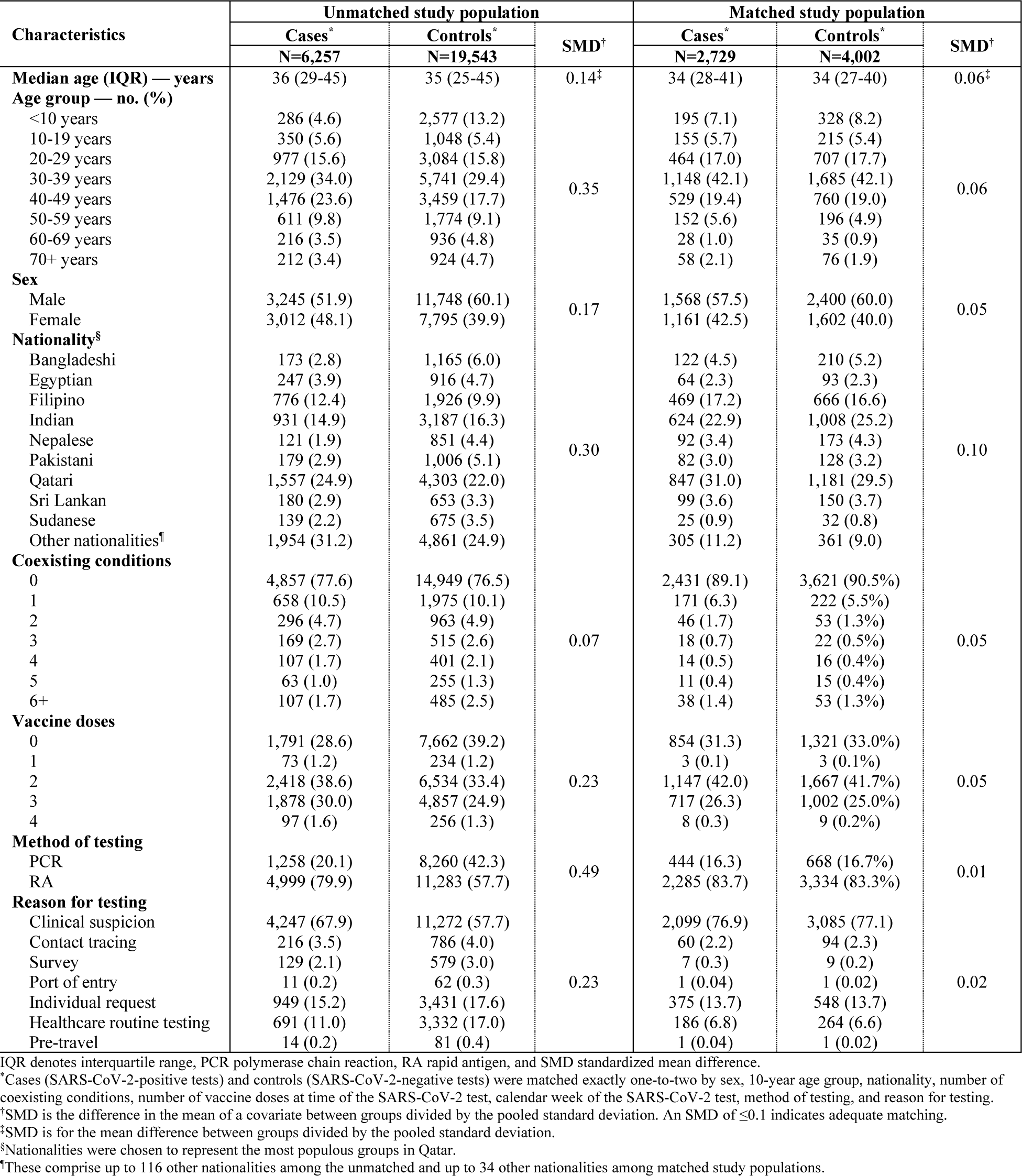
Characteristics of unmatched and matched cases and controls.

**Table S2.** Effectiveness of natural infection in preventing A) symptomatic reinfection with JN.1 and reinfection with JN.1 regardless of symptoms for B) only unvaccinated individuals, and C) only vaccinated individuals.

| Effectiveness of natural infection in preventing | Cases* |  | Controls* |  | Effectiveness† in %<br>(95% CI)‡ |
| --- | --- | --- | --- | --- | --- |
|  | Previous infection (n) | No previous infection (n) | Previous infection (n) | No previous infection (n) |  |
| A) Symptomatic reinfection with JN.1 |  |  |  |  |  |
| Any previous infection | 867 | 1,232 | 1,187 | 1,898 | -2.3 (-14.4 to 10.3) |
| By time since previous infection |  |  |  |  |  |
| Subgroup analysis 1 |  |  |  |  |  |
| 3 months§-<6 months | 2 | 955 | 15 | 1,565 | 82.3 (28.4 to 96.0) |
| 6 months-<9 months | 6 | 956 | 19 | 1,574 | 43.7 (-33.1 to 78.8) |
| 9 months-<1 year | 16 | 961 | 21 | 1,578 | -8.4 (-55.0 to 46.4) |
| ≥1 year | 801 | 1,216 | 1,078 | 1,882 | -4.8 (-16.9 to 8.3) |
| Subgroup analysis 2 |  |  |  |  |  |
| <1 year | 29 | 976 | 60 | 1,590 | 36.9 (-4.8 to 62.1) |
| ≥1 year | 801 | 1,216 | 1,078 | 1,882 | -4.8 (-16.9 to 8.3) |
| B) Reinfection, regardless of symptoms, for only unvaccinated individuals |  |  |  |  |  |
| Any previous infection | 159 | 695 | 257 | 1,064 | 11.8 (-13.2 to 32.4) |
| By time since previous infection |  |  |  |  |  |
| Subgroup analysis 1 |  |  |  |  |  |
| 3 months§-<6 months | 1 | 635 | 8 | 1,003 | 79.3 (-40.3 to 97.4) |
| 6 months-<9 months | 3 | 635 | 6 | 1,006 | -9.6 (-80.2 to 75.8) |
| 9 months-<1 year | 3 | 638 | 9 | 1,005 | 40.8 (-57.0 to 85.0) |
| ≥1 year | 149 | 691 | 228 | 1,060 | 6.9 (-18.8 to 29.6) |
| Subgroup analysis 2 |  |  |  |  |  |
| <1 year | 8 | 640 | 26 | 1,010 | 45.0 (-24.4 to 77.1) |
| ≥1 year | 149 | 691 | 228 | 1,060 | 6.9 (-18.8 to 29.6) |
| C) Reinfection, regardless of symptoms, for only vaccinated individuals |  |  |  |  |  |
| Any previous infection | 999 | 876 | 1,377 | 1,304 | -0.7 (-12.8 to 11.5) |
| By time since previous infection |  |  |  |  |  |
| Subgroup analysis 1 |  |  |  |  |  |
| 3 months§-<6 months | 2 | 560 | 16 | 922 | 83.6 (28.0 to 96.3) |
| 6 months-<9 months | 5 | 562 | 20 | 928 | 64.0 (0.0 to 87.0) |
| 9 months-<1 year | 16 | 565 | 26 | 939 | 10.4 (-45.2 to 56.0) |
| ≥1 year | 917 | 854 | 1,240 | 1,292 | -4.6 (-16.5 to 8.2) |
| Subgroup analysis 2 |  |  |  |  |  |
| <1 year | 29 | 587 | 70 | 948 | 50.5 (16.7 to 70.5) |
| ≥1 year | 917 | 854 | 1,240 | 1,292 | -4.6 (-16.5 to 8.2) |

CI denotes confidence interval, PCR polymerase chain reaction, RA rapid antigen, SARS-CoV-2 severe acute respiratory syndrome coronavirus 2.
\*Cases (SARS-CoV-2-positive tests) and controls (SARS-CoV-2-negative tests) were matched exactly one-to-two by sex, 10-year age group, nationality, number of coexisting conditions, number of vaccine doses at time of the SARS-CoV-2 test, calendar week of the SARS-CoV-2 test, method of testing, and reason for testing.
<sup>†</sup>Effectiveness of previous infection in preventing reinfection was estimated using the test-negative, case-control study design.<sup>27</sup>
<sup>‡</sup>CIs were not adjusted for multiplicity and thus should not be used to infer definitive differences between different groups.
<sup>§</sup>Individuals with a record for a SARS-CoV-2 test within 90 days were excluded from the analysis.

**Table S3.** Strengthening the Reporting of Observational Studies in Epidemiology (STROBE) checklist for case-control studies.

|  | Item No | Recommendation | Main text page |
| --- | --- | --- | --- |
| Title and abstract | 1 | (a) Indicate the study’s design with a commonly used term in the title or the abstract | Manuscript, paragraph 1 |
|  |  | (b) Provide in the abstract an informative and balanced summary of what was done and what was found | Not applicable |
| Introduction |  |  |  |
| Background/rationale | 2 | Explain the scientific background and rationale for the investigation being reported | Manuscript, paragraph 1 |
| Objectives | 3 | State specific objectives, including any prespecified hypotheses | Manuscript, paragraph 1 |
| Methods |  |  |  |
| Study design | 4 | Present key elements of study design | Manuscript, paragraphs 2-3 & Section S3 (‘Study population and study design’) in Supplementary Appendix |
| Setting | 5 | Describe the setting, locations, and relevant dates, including periods of recruitment, exposure, follow-up, and data collection | Manuscript, paragraphs 2-3, & Section S3 (‘Study population and study design’) & Figures S1-S2 in Supplementary Appendix |
| Participants | 6 | (a) Give the eligibility criteria, and the sources and methods of case ascertainment and control selection. Give the rationale for the choice of cases and controls<br>(b) For matched studies, give matching criteria and the number of controls per case | Manuscript, paragraphs 2-3 & Sections S1 & S3 (‘Study population and study design’), & Figure S2 in Supplementary Appendix |
| Variables | 7 | Clearly define all outcomes, exposures, predictors, potential confounders, and effect modifiers. Give diagnostic criteria, if applicable | Manuscript, paragraphs 1 & 3, & Sections S1, S3, & S3 (‘Study population and study design’ & ‘Statistical analysis’), & Table S1 in Supplementary Appendix |
| Data sources/measurement | 8 | For each variable of interest, give sources of data and details of methods of assessment (measurement). Describe comparability of assessment methods if there is more than one group | Manuscript, paragraph 2, & Section S1 in Supplementary Appendix |
| Bias | 9 | Describe any efforts to address potential sources of bias | Manuscript, paragraph 3 & Section S3 (‘Statistical analysis’) in Supplementary Appendix |
| Study size | 10 | Explain how the study size was arrived at | Figure S2 in Supplementary Appendix |
| Quantitative variables | 11 | Explain how quantitative variables were handled in the analyses. If applicable, describe which groupings were chosen and why | Section S3 (‘Study population and study design’) & Table S1 in Supplementary Appendix |
| Statistical methods | 12 | (a) Describe all statistical methods, including those used to control for confounding | Section S3 (‘Statistical analysis’) in Supplementary Appendix |
|  |  | (b) Describe any methods used to examine subgroups and interactions | Section S3 (‘Statistical analysis’) in Supplementary Appendix |
|  |  | (c) Explain how missing data were addressed | Not applicable, see Section S1 in Supplementary Appendix |
|  |  | (d) If applicable, explain how matching of cases and controls was addressed | Manuscript, paragraph 3 & Section S3 (‘Study population and study design’ in Supplementary Appendix |
|  |  | (e) Describe any sensitivity analyses | Section S3 (‘Statistical analysis’) in Supplementary Appendix |
| Results |  |  |  |
| Participants | 13 | (a) Report numbers of individuals at each stage of study—eg numbers potentially eligible, examined for eligibility, confirmed eligible, included in the study, completing follow-up, and analysed<br>(b) Give reasons for non-participation at each stage<br>(c) Consider use of a flow diagram | Figure S2 & Table S1 in Supplementary Appendix |
| Descriptive data | 14 | (a) Give characteristics of study participants (eg demographic, clinical, social) and information on exposures and potential confounders | Tables S1 & S2 in Supplementary Appendix |
|  |  | (b) Indicate number of participants with missing data for each variable of interest | Not applicable, see Section S1 in Supplementary Appendix |
| Outcome data | 15 | Report numbers in each exposure category, or summary measures of exposure | Figure 1 |
| Main results | 16 | (a) Give unadjusted estimates and, if applicable, confounder-adjusted estimates and their precision (eg, 95% confidence interval). Make clear which confounders were adjusted for and why they were included | Manuscript, paragraph 5 & Figure 1 |
|  |  | (b) Report category boundaries when continuous variables were categorized | Table S1 in Supplementary Appendix |
|  |  | (c) If relevant, consider translating estimates of relative risk into absolute risk for a meaningful time period | Not applicable |
| Other analyses | 17 | Report other analyses done—eg analyses of subgroups and interactions, and sensitivity analyses | Manuscript, paragraph 6, Figure 1, & Table S3 in Supplementary Appendix |
| **Discussion** |  |  |  |
| Key results | 18 | Summarise key results with reference to study objectives | Manuscript, paragraph 7 |
| Limitations | 19 | Discuss limitations of the study, taking into account sources of potential bias or imprecision. Discuss both direction and magnitude of any potential bias | Section S3 ('Limitations') in Supplementary Appendix |
| Interpretation | 20 | Give a cautious overall interpretation of results considering objectives, limitations, multiplicity of analyses, results from similar studies, and other relevant evidence | Manuscript, paragraph 7 |
| Generalisability | 21 | Discuss the generalisability (external validity) of the study results | Section S3 ('Limitations') in Supplementary Appendix |
| **Other information** |  |  |  |
| Funding | 22 | Give the source of funding and the role of the funders for the present study and, if applicable, for the original study on which the present article is based | Manuscript, page 6 & Acknowledgements in Supplementary Appendix |

## References

1. Yang S, Yu Y, Xu Y, et al. Fast evolution of SARS-CoV-2 BA.2.86 to JN.1 under heavy immune pressure. Lancet Infect Dis 2024;24:e70–e2.

2. World Health Organization. Excutive Summary Initial Risk Evaluation of JN.1. Available from: https://www.who.int/docs/default-source/coronaviruse/18122023_jn.1_ire_clean.pdf?sfvrsn=6103754a_3. Accessed on January 31, 2024. 2023.

3. Ayoub HH, Tomy M, Chemaitelly H, et al. Estimating protection afforded by prior infection in preventing reinfection: Applying the test-negative study design. Am J Epidemiol 2023.

4. Chemaitelly H, Tang P, Coyle P, et al. Protection against Reinfection with the Omicron BA.2.75 Subvariant. N Engl J Med 2023;388:665–7.

5. Chemaitelly H, Nagelkerke N, Ayoub HH, et al. Duration of immune protection of SARS-CoV-2 natural infection against reinfection. J Travel Med 2022;29.

## References

1. Chemaitelly H, Tang P, Hasan MR, et al. Waning of BNT162b2 Vaccine Protection against SARS-CoV-2 Infection in Qatar. N Engl J Med 2021;385:e83.

2. Altarawneh HN, Chemaitelly H, Ayoub HH, et al. Effects of Previous Infection and Vaccination on Symptomatic Omicron Infections. N Engl J Med 2022;387:21–34.

3. Chemaitelly H, Ayoub HH, AlMukdad S, et al. Bivalent mRNA-1273.214 vaccine effectiveness against SARS-CoV-2 omicron XBB* infections. J Travel Med 2023;30.

4. Altarawneh HN, Chemaitelly H, Hasan MR, et al. Protection against the Omicron Variant from Previous SARS-CoV-2 Infection. N Engl J Med 2022;386:1288–90.

5. Chemaitelly H, Ayoub HH, Tang P, et al. Long-term COVID-19 booster effectiveness by infection history and clinical vulnerability and immune imprinting: a retrospective population-based cohort study. Lancet Infect Dis 2023;23:816–27.

6. Abu-Raddad LJ, Chemaitelly H, Bertollini R, National Study Group for Covid Vaccination. Effectiveness of mRNA-1273 and BNT162b2 Vaccines in Qatar. N Engl J Med 2022;386:799–800.

7. Al-Thani MH, Farag E, Bertollini R, et al. SARS-CoV-2 Infection Is at Herd Immunity in the Majority Segment of the Population of Qatar. Open Forum Infect Dis 2021;8:ofab221.

8. Planning and Statistics Authority-State of Qatar. Qatar Monthly Statistics. Available from: https://www.psa.gov.qa/en/pages/default.aspx. Accessed on: May 26, 2020. 2020.

9. Abu-Raddad LJ, Chemaitelly H, Ayoub HH, et al. Characterizing the Qatar advanced-phase SARS-CoV-2 epidemic. Sci Rep 2021;11:6233.

10. Chemaitelly H, Bertollini R, Abu-Raddad LJ, National Study Group for Covid Epidemiology. Efficacy of Natural Immunity against SARS-CoV-2 Reinfection with the Beta Variant. N Engl J Med 2021;385:2585–6.

11. Abu-Raddad LJ, Chemaitelly H, Ayoub HH, et al. Effect of mRNA Vaccine Boosters against SARS-CoV-2 Omicron Infection in Qatar. N Engl J Med 2022;386:1804–16.

12. Chemaitelly H, Faust JS, Krumholz HM, et al. Short- and longer-term all-cause mortality among SARS-CoV-2-infected individuals and the pull-forward phenomenon in Qatar: a national cohort study. Int J Infect Dis 2023;136:81–90.

13. AlNuaimi AA, Chemaitelly H, Semaan S, et al. All-cause and COVID-19 mortality in Qatar during the COVID-19 pandemic. BMJ Glob Health 2023;8.

14. Vogels C, Fauver J, Grubaugh N. Multiplexed RT-qPCR to screen for SARS-COV-2 B.1.1.7, B.1.351, and P.1 variants of concern V.3. 10.17504/protocols.io.br9vm966. 2021.

15. Abu-Raddad LJ, Chemaitelly H, Butt AA, National Study Group for Covid Vaccination. Effectiveness of the BNT162b2 Covid-19 Vaccine against the B.1.1.7 and B.1.351 Variants. N Engl J Med 2021;385:187–9.

16. Chemaitelly H, Yassine HM, Benslimane FM, et al. mRNA-1273 COVID-19 vaccine effectiveness against the B.1.1.7 and B.1.351 variants and severe COVID-19 disease in Qatar. Nat Med 2021;27:1614–21.

17. Qatar viral genome sequencing data. Data on randomly collected samples. https://www.gisaid.org/phylodynamics/global/nextstrain/. 2021. at https://www.gisaid.org/phylodynamics/global/nextstrain/.)

18. Benslimane FM, Al Khatib HA, Al-Jamal O, et al. One Year of SARS-CoV-2: Genomic Characterization of COVID-19 Outbreak in Qatar. Front Cell Infect Microbiol 2021;11:768883.

19. Hasan MR, Kalikiri MKR, Mirza F, et al. Real-Time SARS-CoV-2 Genotyping by High-Throughput Multiplex PCR Reveals the Epidemiology of the Variants of Concern in Qatar. Int J Infect Dis 2021;112:52–4.

20. Saththasivam J, El-Malah SS, Gomez TA, et al. COVID-19 (SARS-CoV-2) outbreak monitoring using wastewater-based epidemiology in Qatar. Sci Total Environ 2021;774:145608.

21. El-Malah SS, Saththasivam J, Jabbar KA, et al. Application of human RNase P normalization for the realistic estimation of SARS-CoV-2 viral load in wastewater: A perspective from Qatar wastewater surveillance. Environ Technol Innov 2022;27:102775.

22. Tang P, Hasan MR, Chemaitelly H, et al. BNT162b2 and mRNA-1273 COVID-19 vaccine effectiveness against the SARS-CoV-2 Delta variant in Qatar. Nat Med 2021;27:2136–43.

23. Chemaitelly H, Ayoub HH, AlMukdad S, et al. Duration of mRNA vaccine protection against SARS-CoV-2 Omicron BA.1 and BA.2 subvariants in Qatar. Nat Commun 2022;13:3082.

24. Qassim SH, Chemaitelly H, Ayoub HH, et al. Effects of BA.1/BA.2 subvariant, vaccination and prior infection on infectiousness of SARS-CoV-2 omicron infections. J Travel Med 2022;29.

25. Altarawneh HN, Chemaitelly H, Ayoub HH, et al. Protective Effect of Previous SARS-CoV-2 Infection against Omicron BA.4 and BA.5 Subvariants. N Engl J Med 2022;387:1620–2.

26. Chemaitelly H, Tang P, Coyle P, et al. Protection against Reinfection with the Omicron BA.2.75 Subvariant. N Engl J Med 2023;388:665–7.

27. Ayoub HH, Tomy M, Chemaitelly H, et al. Estimating protection afforded by prior infection in preventing reinfection: Applying the test-negative study design. Am J Epidemiol 2023.

28. Altarawneh HN, Chemaitelly H, Ayoub HH, et al. Effects of previous infection, vaccination, and hybrid immunity against symptomatic Alpha, Beta, and Delta SARS-CoV-2 infections: an observational study. EBioMedicine 2023;95:104734.

29. Qassim SH, Chemaitelly H, Ayoub HH, et al. Population immunity of natural infection, primary-series vaccination, and booster vaccination in Qatar during the COVID-19 pandemic: an observational study. eClinicalMedicine 2023;62:102102.

30. Jackson ML, Nelson JC. The test-negative design for estimating influenza vaccine effectiveness. Vaccine 2013;31:2165–8.

31. Verani JR, Baqui AH, Broome CV, et al. Case-control vaccine effectiveness studies: Preparation, design, and enrollment of cases and controls. Vaccine 2017;35:3295–302.

32. Pilz S, Theiler-Schwetz V, Trummer C, Krause R, Ioannidis JPA. SARS-CoV-2 reinfections: Overview of efficacy and duration of natural and hybrid immunity. Environ Res 2022:112911.

33. Kojima N, Shrestha NK, Klausner JD. A Systematic Review of the Protective Effect of Prior SARS-CoV-2 Infection on Repeat Infection. Eval Health Prof 2021;44:327–32.

34. Ayoub HH, Chemaitelly H, Seedat S, et al. Mathematical modeling of the SARS-CoV-2 epidemic in Qatar and its impact on the national response to COVID-19. J Glob Health 2021;11:05005.

35. Coyle PV, Chemaitelly H, Ben Hadj Kacem MA, et al. SARS-CoV-2 seroprevalence in the urban population of Qatar: An analysis of antibody testing on a sample of 112,941 individuals. iScience 2021;24:102646.

36. Jeremijenko A, Chemaitelly H, Ayoub HH, et al. Herd Immunity against Severe Acute Respiratory Syndrome Coronavirus 2 Infection in 10 Communities, Qatar. Emerg Infect Dis 2021;27:1343–52.

37. Abu-Raddad LJ, Chemaitelly H, Bertollini R, National Study Group for Covid Vaccination. Waning mRNA-1273 Vaccine Effectiveness against SARS-CoV-2 Infection in Qatar. N Engl J Med 2022;386:1091–3.

38. Chemaitelly H, Ayoub HH, Coyle P, et al. Protection of Omicron sub-lineage infection against reinfection with another Omicron sub-lineage. Nat Commun 2022;13:4675.

39. Austin PC. Using the Standardized Difference to Compare the Prevalence of a Binary Variable Between Two Groups in Observational Research. Communications in Statistics - Simulation and Computation 2009;38:1228–34.

40. Tseng HF, Ackerson BK, Bruxvoort KJ, et al. Effectiveness of mRNA-1273 vaccination against SARS-CoV-2 omicron subvariants BA.1, BA.2, BA.2.12.1, BA.4, and BA.5. Nature Communications 2023;14:189.

41. Abu-Raddad LJ, Chemaitelly H, Yassine HM, et al. Pfizer-BioNTech mRNA BNT162b2 Covid-19 vaccine protection against variants of concern after one versus two doses. J Travel Med 2021;28.

